# Decision curve analysis to identify optimal candidates of liver resection for intermediate-stage hepatocellular carcinoma with hepatitis B cirrhosis : A cohort study

**DOI:** 10.1101/2020.10.07.20166769

**Authors:** Siyu Chen, Linbin Lu, Xinyu Hu, Shan Lin, Lijun Zhu

## Abstract

**Background:** The selection criterion for liver resection (LR) in intermediate-stage (IM) hepatocellular carcinoma (HCC) is still controversial. This study aims to compare LR and transarterial chemoembolization in the range of predicted death risk

**Methods:** The multivariable Cox regression model (MVR) was estimated to predict mortality at 5yr. The cut-off values were determined by a two-piece-wise linear regression model, decision curve analysis with MVR model, and hazard ratio curve for treatment plotted against the predicted mortality.

**Results:** 825 IM-HCC with hepatitis B cirrhosis were included for analysis (TACE, n=622; LR, n=203). The 5-yr overall survival rate of LR patients was higher than the TACE group (52.8% vs. 20.8%; *P*<0.0001). The line of LR and TACE were crossing with predicted death risk at 100% (*P* for interaction =0.008). The benefit of LR versus TACE decreased progressively as predicted death risk>0.55 (95%CI: 0.45,0.62). When predicted death risk over 0.7, decision curve analysis suggested that LR and TACE did not increase net benefit. Patients were then divided into four subgroups by the cut-off values (<0.45, 0.45≥ /<0.62, 0.62≥ /<0.7, ≥ 0.7). The stratified analysis of treatment in different subgroups, hazard ratios were 0.39 (95%CI: 0.27, 0.56), 0.36 (95%CI: 0.23, 0.56), 0.51 (95%CI: 0.27, 0.98), and 0.46 (95%CI: 0.27, 0.80), respectively.

**Conclusions:** LR reached the maximal relative utility in the interval of 0.45 to 0.62, and both LR and TACE did not increase net benefit at the 5-yr death risk over 0.7.

## 1. Introduction

Hepatocellular carcinoma (HCC) is the fifth cause of death in China, and its high levels of morbidity and mortality rates mostly result from hepatitis B cirrhosis, with nearly half of the global new cases and deaths^1,2^. Liver resection, regarded as a curative therapy, is widely performed for the patients with Barcelona Clinic Liver Cancer (BCLC) 0-A stage HCC in China, even for the resectable intermediate-stage tumor^3^. According to the last BCLC staging system, transarterial chemoembolization (TACE) has been recommended as the first-line treatment for unresectable intermediate-stage(IM) HCC^4^. In the clinical setting, however, transarterial chemoembolization (TACE) is also applied on a wide scale in resectable HCC patients alone or combined with surgery. The advantage of surgery over TACE has already been extensively described in the resectable BCLC-B HCC patients ^5^ and the cirrhotic patients^6^. However, which groups are more suitable for liver resection (LR) remains an open issue.

Recently, an NSP scoring system (1, >1 point; median OS, 61.3 vs. 19.3 months) was developed to select patients with IM-HCC for LR accurately^7^. Compared with the TACE treatment, surgical resection has the better survival only in the subclass of BCLC-B HCC with a Child-Pugh score of 5 and no more than 3 tumors^8^. Besides, according to the modified Bolondi’s sub-staging model, partial hepatectomy has an optimal long-term survival than TACE in the BCLC-B1/B2 HCC^9^. Interestingly, in another study, patients were grouped into low-risk (3yr mortality ≤ 35%), medium-risk (35%< 3yr mortality < 70%), and high-risk (3yr mortality ≥ 70%) groups by regret-based decision curve analysis. And only the patients in the low-risk group have a better outcome than those treated with TACE^10^. However, more convenient biomarkers are urgent to choose suitable therapy for IM-HCC, a highly heterogeneous population. In the present study, we use a predicted mortality risk-based decision analysis to compare the clinical outcome of hepatectomy and TACE in the IM-HCC. We present the following article in accordance with the STROBE reporting checklist.

## 2. Material and Methods

### 2.1 Patient selection

Clinical and biological data have been previously published in full^11,12^, and the details of inclusion criteria, diagnosis, and treatment are shown in Figure S1. The Ethics Committee approved the study protocol (2017-FXY-129) of SYSUCC and another three medical centers^11^. Because this was a retrospective study, the informed consent was waived.

### 2.2 Definition and follow up

Liver resection (LR): surgical therapy for hepatic segments or lobes lesions. In the clinical, patients with well liver function and less tumor loading were usually suitable for HR.

Transarterial chemoembolization (TACE): chemoembolization of the hepatic artery. According to the response to the TACE, the subsequent therapies include ablative therapies, surgical resection, TACE, or targeted therapies.

Overall survival (OS): the time between the beginning of LR/TACE and the last following-up or death for any cause.

During the initial treatment period for the first 2 years, patients were followed up for every 2 or 3 months if complete remission was achieved. The frequency gradually decreased to every 3 to 6 months after remission of 2 years.

### 3.3 Statistical analyses

To compare differences of baseline characteristics between the HR and TACE groups, we compared categorical variables with the chi-square test and continuous variables by the Mann-Whitney test. Kaplan-Meier methods calculated overall survival rates for the LR and TACE cohorts. Statistical analysis was performed using Empower (www.empowerstats.com, X&Y solutions, inc. Boston MA) and R software (version 3.4.3). P-value < 0.05 considered significant.

To explore the optimal range of death risk for the net benefit of LR against TACE, we divide the statistical analyses into three steps:

Firstly, according to the previously reported methods^13^, we developed the multivariable Cox regression (MVR) model, which was based on the significantly different covariates between two treatment groups (all *P*<0.05 list in Table 1), including the continuous covariates of age, PT and diameter of the main tumor, as well as the categorical covariates *No*. of intrahepatic lesions, both lobe with lesions.

**Table 1.** Baseline characteristics between TACE and surgery group in the derivation cohort.

|  | Treatment |  | Standardize difference | P-value |
| --- | --- | --- | --- | --- |
|  | TACE | Surgery |  |  |
| No. | 622 | 203 |  |  |
| Age | 53.9 ± 12.2 | 50.6 ± 12.4 | 0.3 (0.1, 0.4) | 0.001 |
| Gender |  |  | 0.0 (-0.1, 0.2) | 0.716 |
| male | 566 (91.0%) | 183 (90.1%) |  |  |
| female | 56 (9.0%) | 20 (9.9%) |  |  |
| AFP (ng/ml) Log | 2.6 ± 1.4 | 2.4 ± 1.5 | 0.2 (-0.0, 0.3) | 0.050 |
| PT (seconds) | 12.3 ± 1.3 | 12.5 ± 1.6 | 0.2 (0.0, 0.3) | 0.037 |
| ALB (g/L) | 39.0 ± 5.8 | 38.4 ± 5.4 | 0.1 (-0.1, 0.3) | 0.220 |
| Log TBLT(umol/L) | 1.3 ± 0.2 | 1.3 ± 0.3 | 0.1 (-0.0, 0.3) | 0.157 |
| Major tumor size(mm) | 75.2 ± 37.0 | 63.6 ± 27.6 | 0.4 (0.2, 0.5) | <0.001 |
| No. of intrahepatic lesions |  |  | 0.7 (0.5, 0.9) | <0.001 |
| 2 | 150 (24.1%) | 110 (54.2%) |  |  |
| 3 | 51 (8.2%) | 20 (9.9%) |  |  |
| >3 | 421 (67.7%) | 73 (36.0%) |  |  |
| Location of Lesions |  |  | 0.6 (0.5, 0.8) | <0.001 |
| 0 | 219 (35.2%) | 132 (65.0%) |  |  |
| 1 | 403 (64.8%) | 71 (35.0%) |  |  |
| Child-Pugh class |  |  | 0.1 (-0.1, 0.3) | 0.246 |
| A | 522 (86.9%) | 168 (83.6%) |  |  |
| B | 79 (13.1%) | 33 (16.4%) |  |  |

Numbers that do not add up to 825 are attributable to missing data.
AFP=alpha-fetoprotein, PT=prothrombin time, TBLT=total bilirubin, ALB=albumin,TACE=transarterial chemoembolisation. The chi-square test was performed for categorical measures and Kruskal-Wallis Test for continuous measures.

To evaluate the interaction between LR and death risk, we used the method^13^: (1) predicted OS rate at 5 yr (Pi) was calculated based on the MVR model in the LR group. From this model, death risk at 5 yr(1-Pi) was also the baseline death risk for both LR and TACE cohorts. (2) this predicted death risk (1-Pi) was added as a covariate to the second multivariate Cox model to calculate the predicted probability of survival at 5 yr (Pii), and (3) spline smoothing curve between 5yr predicted death risk (1-Pi) and probability (Pii) were graphed stratified by treatment through the generalized additive model. Furthermore, a two-piece-wise linear regression and the recursive method were performed to calculate the inflexion point of the TACE line, and a log-likelihood ratio test was used to compare the one-line linear regression. The 95% confidence interval of inflection point was confirmed by 500 bootstrap resampling, which may be the optimal range for the net benefit of LR against TACE.

Secondly, to further verify this interval, we used the “ggDCA” R package to establish a decision curve analysis with the MVR model. We calculated the net benefit of the model and determined the cut-off value through two reference strategies (test none or test all).

Thirdly, to further validate the cut-off points from the two steps above, we paint the hazard ratio (HR) curve (LR vs. TACE) for the 5yr predicted death risk by the Cox regression model. The stratified analyses were performed to explore LR (vs. TACE) hazard ratio in each subgroup based on the observed cut-off values.

## 3. Results

### 3.1 Descriptive characteristics

In the derivation cohort, 825 patients with hepatitis B cirrhosis were included in the final analysis, with TACE (n=622, 75.4%) or surgical resection (n=203, 24.6%) as their first-line treatment. Table 1 showed the baseline features of study populations. The univariable analysis results were listed in Table S1, focusing on the markedly different covariates between the two groups.

The median overall survival for the entire cohort was 23.3 (95%CI: 20.0, 26.6) month. And it was 18.0(95%:16.2,19.9) month for TACE group versus 67.4(95%CI:44.0, NA) month for LR group (*P*<0.0001, see Figure S2). The OS rates at 5 yr was 29.5% (95%CI: 25.7%, 33.9%) for the entire cohort, and it was 52.8% (95% CI:45.0%, 62.0%) for HR group versus 20.8% (95%CI: 16.8%, 25.7%) for the TACE group (*P* < 0.0001).

### 3.2 Survival analysis for net benefit in treatment cohorts

The MVR model was estimated by age, PT, largest tumor size, *No*. of intrahepatic lesions, and both lobes with lesions, which was used to predict 5-yr death risk and OS rate for the entire cohort (Figure S3). Its Harrell’s C index was 0.70 (0.67, 0.72). In Figure 1, the predicted death risk was plotted against OS rate at 5yr. And the lines for LR and TACE crossed at 1.0 (*P* for interaction = 0.008). In Table 2, we found that the inflexion point of the TACE line was calculated at 0.55 (95%CI: 0.45, 0.62), indicating the benefit from HR decreased progressively as predicted OM risk over 55%. Interestingly, the predicted survival rate of the TACE line at 0.55 was 0.20 (95%CI: 0.18, 0.21), which was in the interval of observed OS at 5yr for the TACE group (20.8%, 95%CI: 16.8%, 25.7%).

**Figure 1.**
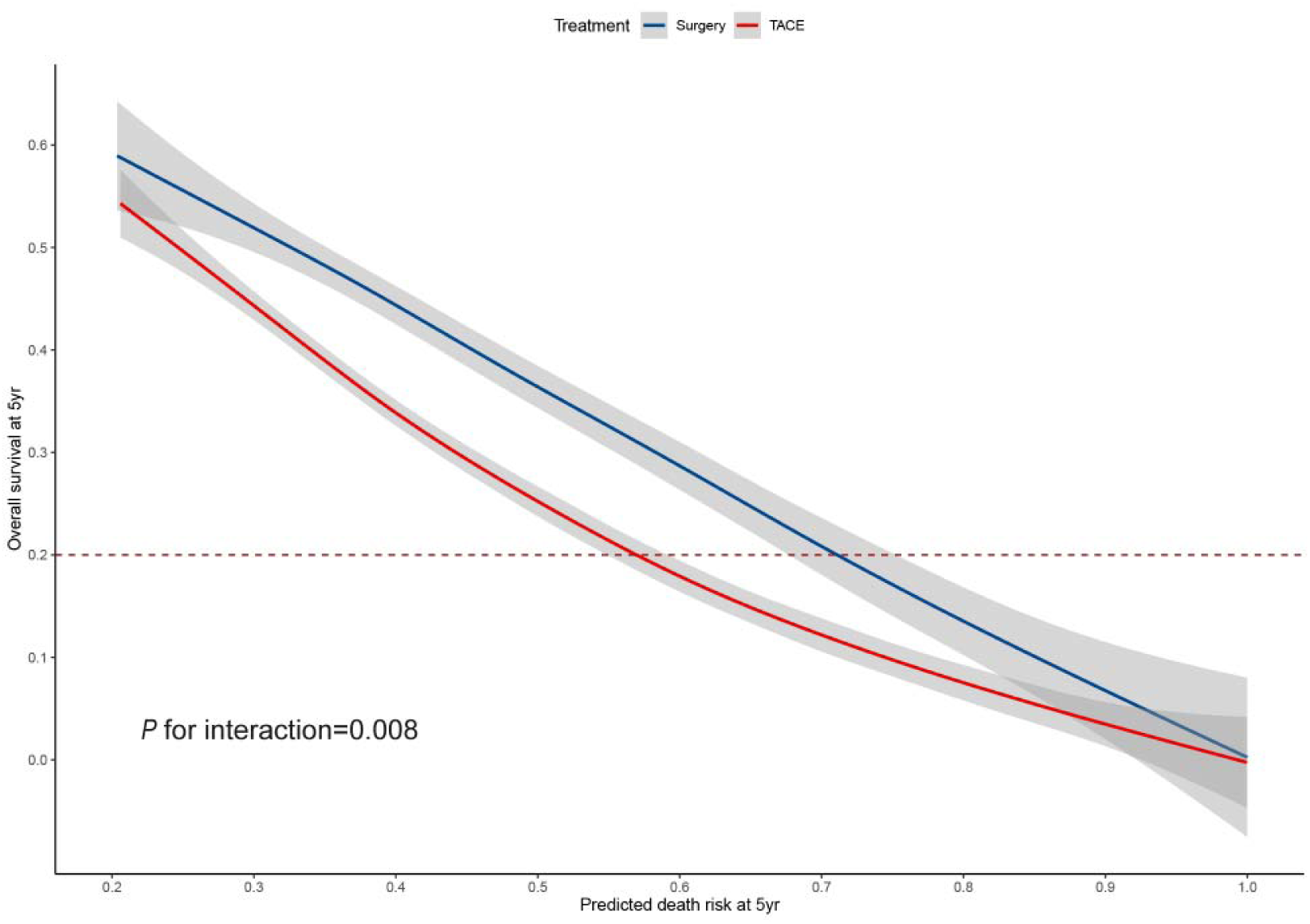
Overall survival rate plotted against predicted probability of death at 5yr. The blue line indicates hepatic resection for primary treatment. The red line indicates TACE for primary treatment. Dashed line indicates observed OS at 5yr for the TACE group (20.8%, 95%CI: 16.8%, 25.7%). The inflexion point of the TACE line was calculated at 0.55 (95%CI: 0.45, 0.62).

**Table 2.** Threshold effect analysis of TACE group in the derivation cohort using two-piece-wise linear regression.

| | Unadjusted $\beta$(SD) | P-value |
| --- | --- | --- |
| The one-line linear model | -0.71(0.02) | <0.0001 |
| The two-piece-wise linear model |  |  |
| < 0.55 | -0.98(0.05) | <0.0001 |
| > 0.55 | -0.47(0.04) | <0.0001 |
| *P* for log-likelihood ratio test |  | <0.001 |

The predicted value at the point of 0.55(95%CI: 0.45, 0.62) was 0.20 (95%CI: 0.18, 0.21). A log-likelihood ratio test was used to compare the one-line linear regression.

As was shown in Figure 2, net benefit curves suggested none could receive a net benefit from surgery and TACE for death risk >0.7; maximal relative utility occurred at 0.4. Consistently, these cut-off values were further validated by the hazard ratio curve of LR and TACE against the 5yr predicted death risk, which was shown in Figure 3. Thus, patients were grouped into four subclasses: R5<0.45, 0.45≥ R5<0.62, 0.62≥ R5<0.7, R5≥ 0.7. Figure 3 also illustrated the stratified analysis of LR (vs TACE), hazard ratios were 0.39 (95%CI: 0.27, 0.56), 0.36 (95%CI: 0.23, 0.56), 0.51 (95%CI: 0.27, 0.98), and 0.46 (95%CI: 0.27, 0.80) for each subclass.

**Figure 2.**
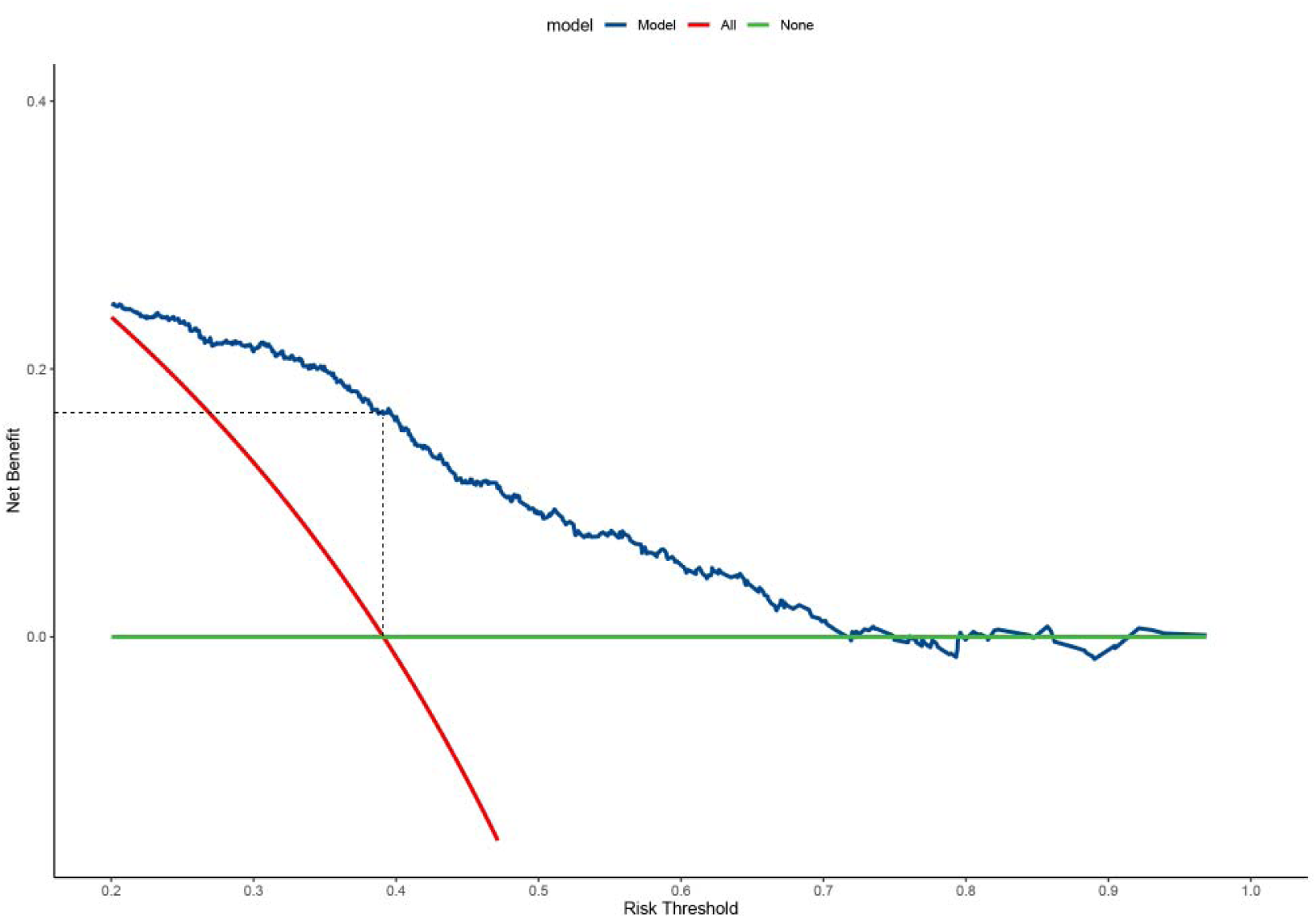
Decision curve analysis with multivariable Cox regression model for overall survival. Solid green line = net benefit when all BCLC-B HCC is considered not having the outcome; red dashed line=net benefit when all pregnant women are considered the outcome.

**Figure 3.**
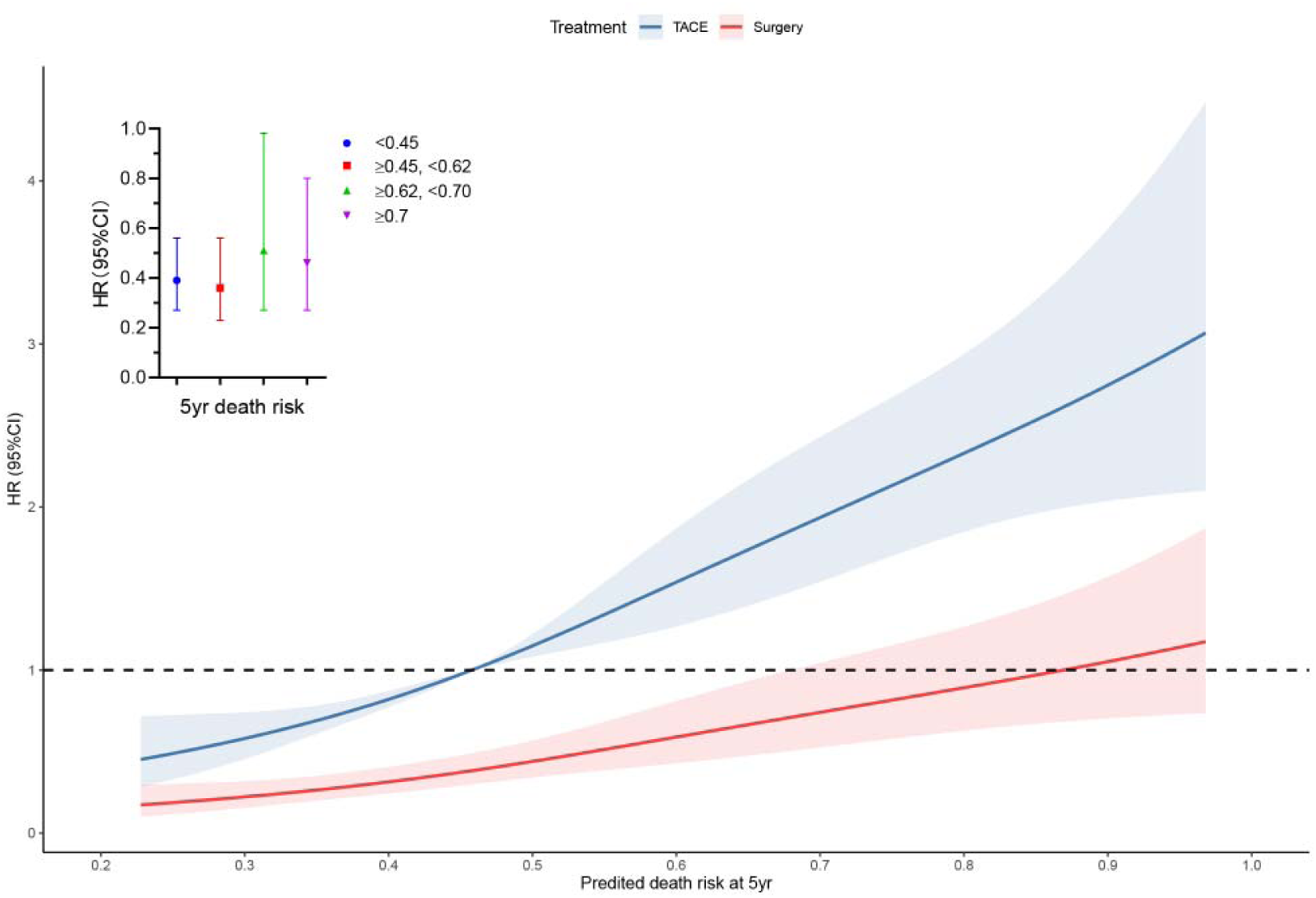
Hazard ratio of treatment plotted against the predicted mortality at 5yr. Hazard ratios for LR versus TACE in the stratified analysis are 0.39 (95%CI: 0.27, 0.56), 0.36 (95%CI: 0.23, 0.56), 0.51 (95%CI: 0.27, 0.98), and 0.46 (95%CI: 0.27, 0.80) for each subclass.

## 4. Discussion

In this large-scale, real-world data, we found that the overall survival for LR was significantly better than their TACE counterparts, which was consistent with the previous literature^5,14^. However, the net benefit from LR decreased progressively as predicted death risk >55%, with the interval of maximal relative utility ranging from 45% to 62%. When death risk>70%, LR and TACE did not increase net benefit.

In 2015, Colombo et al. ^15^ had come up with an assumption that intermediate-stage HCC patients could still be suitable for liver resection if the 5-yr survival rate reached 50%. Our findings primarily validated this hypothesis. In line with the previous literature ^9,16^, we identified a subgroup with death risk<70%, in which patients treated with HR had significantly better overall survival than the TACE group. Based on Bolondi’s sub-staging model^17^, Wei et al. ^9^, patients’ postoperative 5-yr survival rate in the BCLC B1-B3 stage was 49.5%, 33.7%, and 12.9%, respectively. Nevertheless, only the BCLC B1/B2 had more optimal long-term survival than the TACE group. In another large-scale study^16^, the benefit from liver resection was observed in the patients of BCLC-B1/B2 but not B3/B4.

Our study had some strengths. We used three novel methods to calculate the cut-off accuracy value to evaluate the tumor loading. Besides, in our study, the survival rates between LR and TACE were compared in the vast and continuous range so that the exact cut-off values could be calculated. When evaluating the role of liver resection among patients with anatomically resectable tumors and well liver function, the randomized control trial was obviously against medical ethics. Therefore, real-world data’s predicted mortality risk-based decision analysis may be better.

Our study also had several limitations. Firstly, this was a retrospective cohort from real-world data; residual bias and unmeasured confounders were unavoidable. Secondly, the percentage of resectable HCC patients in the TACE group with 5-yr death risk< 70% was unclear. However, it was worthy of note that the potential unresectable HCC patients treated with TACE resulting from such errors would bias toward the null and lead to an underestimation of the net benefit from liver resection versus TACE.

## 5. Conclusions

LR reached the maximal relative utility in the interval of 0.45 to 0.62, and both LR and TACE did not increase net benefit at the 5-yr death risk over 0.7.

## Data Availability

The raw data were freely obtained from the Dryad Digital Repository database (www. Datadryad.org; https://doi.org/10.5061/dryad.pd44k8r).

## Abbreviations

BCLC: Barcelona clinic liver cancer
SYSUCC: Sun Yat-sen University Cancer Center
LR: Liver resection
IM-HCC: intermediate-stage hepatocellular carcinoma
TACE: transarterial chemoembolization
OS: overall survival
HBV: hepatitis B virus

## Ethics approval and informed consent

The study protocol (2017-FXY-129) was approved by the clinical research department at SYSUCC. Because this was a secondary analysis study, and the data were anonymous, the requirement for informed consent was waived.

## Data availabilit

The data are available from the corresponding author on reasonable request.

## Author contributions

(I) Conception and design: All authors; (II) Administrative support: All authors; (III) Provision of study materials or patients: All authors; (IV) Collection and assembly of data: All authors; (V) Data analysis and interpretation: All authors; (VI) Manuscript writing: All authors;(VII) Final approval of manuscript: All authors

## Acknowledgments

We gratefully thank you for the raw data from Prof. Peihong Wu and the statistical support from Empower U team of the Department of Epidemiology and Biostatistics, X&Y solutions Inc. Funding:

## Supplemental materials

**Figure S1.**
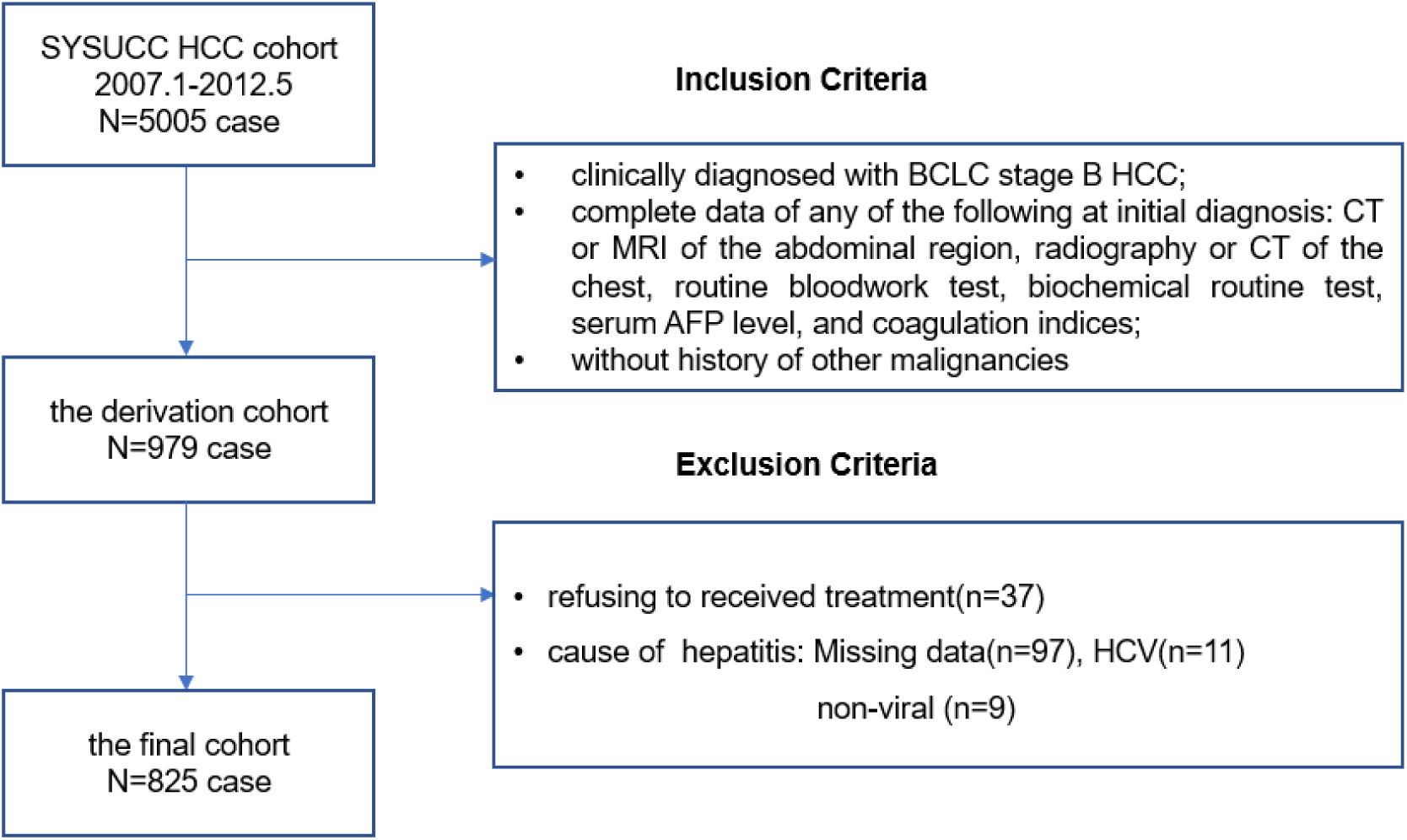
Inclusion and exclusion criteria of hepatocellular carcinoma patients. Between January 2007 and May 2012, 5005 consecutive patients with newly diagnosed HCC at Sun Yat-sen University Cancer Center (SYSUCC) were retrospectively reviewed to develop the derivation cohort. After meeting the inclusion criteria, a total of 979 patients were included in the derivation cohort.

### Diagnosis

According to the exclusion criteria, 825 cases were included in the final cohort. For the patients treated with surgery (n=203), HCC diagnosis was confirmed by histopathological examination of surgical samples. For the patients with TACE(n=622), in contrast, the diagnosis was confirmed by the combination with the serum level of alpha-fetoprotein (AFP, over 400 ng/mL) and clinical imaging, including ultrasonography, computed tomography, or magnetic resonance imaging. A needle biopsy was performed if the diagnosis was uncertain based on imaging and AFP level.

### Treatment

Based on the decisions of the multidisciplinary teams, the optimal treatment plan was adopted for each HCC patient. The indications for surgery in IM-HCC patients were appropriate residual liver volume determined by computed tomography. For patients without cirrhosis, 30% remnant liver volume after HR was considered adequate. However, for those with chronic hepatitis cirrhosis, the remnant volume should be more than 50%. Patients who satisfied the indications for surgery were treated by surgical resection unless the patient requested TACE.

**Figure S2.**
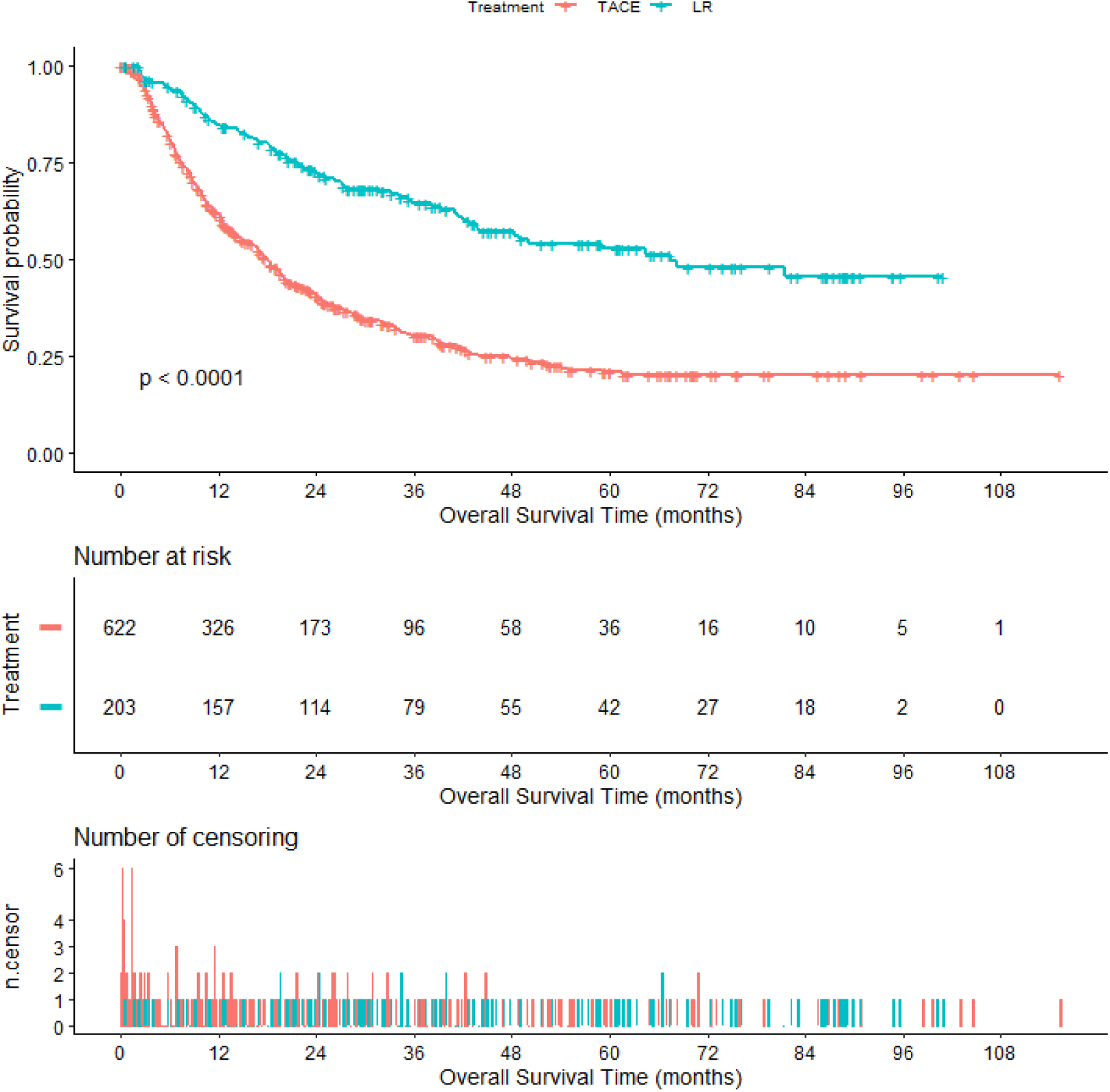
Kaplan-Meier curves of overall survival in the derivation cohort stratified with liver resection and transarterial chemoembolization.

**Figure S3.**
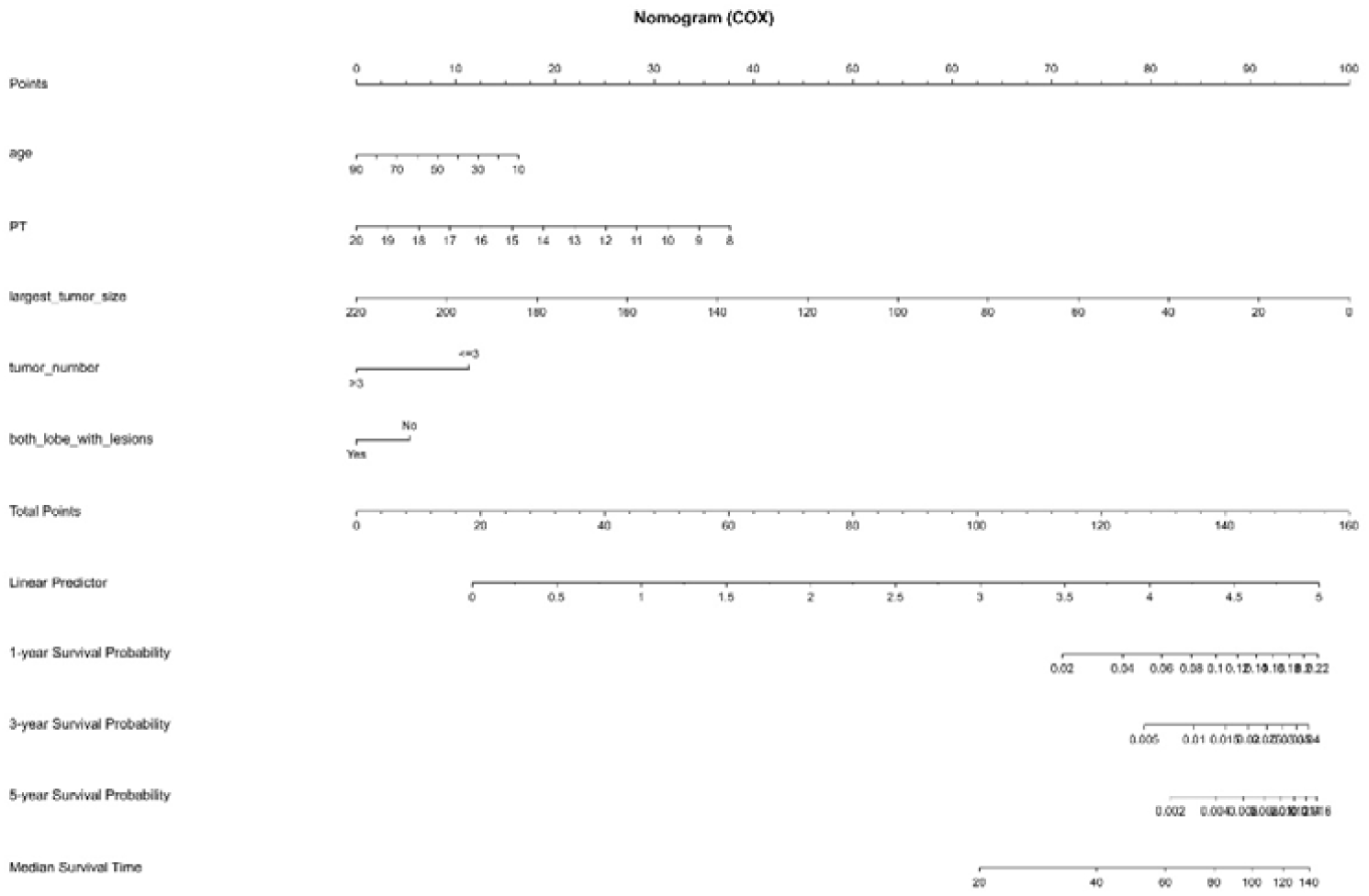
Nomogram to predict the OS. 1yr-,3yr-and 5yr-AUC are 0.73, 0.69, 0.66.

**Table S1.** The univariate analysis focusing on the derivation cohort.

|  | Statistics | Death | P-value |
| --- | --- | --- | --- |
| Treatment |  |  |  |
| TACE | 622 (75.4%) | Reference |  |
| LR | 203 (24.6%) | 0.39 (0.30, 0.50) | <0.0001 |
| Age | 53.2 ± 12.5 | 1.00 (0.99, 1.01) | 0.661 |
| Gender |  |  | 0.381 |
| male | 749 (90.8%) | Reference |  |
| female | 76 (9.2%) | 1.15 (0.84, 1.58) |  |
| ALB (g/L) | 38.8± 5.7 | 0.99 (0.97, 1.00) | 0.104 |
| PT (second) | 12.3 ± 1.4 | 1.09 (1.01, 1.16) | 0.017 |
| No. of intrahepatic lesions |  |  |  |
| 2 | 260 (31.5%) | Reference |  |
| 3 | 71 (8.6%) | 1.11 (0.78, 1.59) | 0.554 |
| >3 | 494 (59.9%) | 1.57 (1.28, 1.94) | <0.0001 |
| Major tumor size(mm) | 72.3 ± 35.3 | 1.01 (1.01, 1.02) | <0.0001 |
| Both lobes with lesion |  |  |  |
| no | 351 (42.5%) | Reference |  |
| yes | 474 (57.5%) | 1.44 (1.19, 1.74) | 0.0001 |
| log<sub>10</sub> AFP (ng/ml) | 2.5 ± 1.4 | 1.21 (1.13, 1.29) | <0.0001 |
| log<sub>10</sub> TBLT(umol/L) | 1.3 ± 0.3 | 0.98 (0.67, 1.43) | 0.909 |
| Child-Pugh class |  |  | 0.009 |
| A | 690 (86.0%) | Reference |  |
| B | 112 (14.0%) | 1.41 (1.09, 1.83) |  |

Numbers that do not add up to 825 are attributable to missing data. AFP=alpha-fetoprotein, PT=prothrombin time, TBLT=total bilirubin, ALB=albumin, HR=hepatic resection.

